# Understanding health insurance coverage and its socio-demographic associations among women in Bangladesh: evidence from BDHS 2022

**DOI:** 10.1101/2025.03.04.25323347

**Authors:** Md Fuad Al Fidah, Md Ridwan Islam, Sharif Mohammad

**Author notes:** **Correspondence:** Dr. Md Fuad Al Fidah, Bibliophile, Dhaka 1212, Bangladesh. **Funding statement:** None received. **Author contributions:** MFAF and SM conceived and designed the study. MFAF and MRI were associated with data collection, curation, analysis and interpretation. MRI and MFAF wrote the manuscript. SM critically reviewed and edited the manuscript. All the author(s) read and approved the final manuscript for submission. **Ethical standards disclosure:** We utilized secondary data obtained from the DHS website (https://dhsprogram.com/) on request, where all identifiable information has been removed to ensure anonymity. As we conducted analysis on secondary data, no ethical clearance is required. **Availability of data:** Data related to this manuscript are available upon request from the DHS website. **Competing interests:** The authors declare that they have no competing interests. **Declaration of Al usage:** The authors used ChatGPT (version 4.o) for proof reading and language editing as the authors are not native English speaker. The authors reviewed and edited the content as needed and takes full responsibility for the content of the publication. **Patient and Public Involvement:** Patients or the public WERE NOT involved in the design, or conduct, or reporting, or dissemination plans of our research.

## Abstract

**Background:** Health insurance is vital for healthcare financing and aligns with the 2030 Sustainable Development Goal agenda. Women of reproductive age face unique barriers to affordable care due to higher health risks and caregiving roles. This study aims to estimate the proportion of women with health insurance coverage (HIC) and its associated factors.

**Methods:** We used data from the latest round of the Bangladesh Demographic and Health Survey (BDHS) of 2022. The number of women considered for analysis was 19987. Unadjusted odds ratio with their95% confidence interval (Cl) were reported.

**Results:** The median (IQR) age of the women in the study was 32.0 (25.0-39.0) years. Only 0.3% of women reported having HIC. The odds of HIC increased significantly with having account with bank or other financial institution (UOR: 2.27; 95% Cl: 1.36-3.79), both woman or their husbands having post secondary education, belonging to richest family (UOR: 3.32; 95% Cl: 1.45-7.60), and having adequate media exposure (UOR: 2.78; 95% Cl: 1.41-5.47).

**Conclusions:** The study highlights the need for higher education among both sexes and targeted strategies to reduce wealth-based disparities and use mass media to promote the uptake of HIC.

**WHAT IS ALREADY KNOWN ON THIS TOPIC:**

- Health insurance is a key mechanism for financing healthcare globally, aligning with the Sustainable Development Goals agenda for 2030.
- Women in reproductive age groups face unique barriers to affordable healthcare, particularly in LMICs.
- Less than 1% of the Bangladeshi population, benefit from social health protection

**WHAT THIS STUDY ADDS:**

- Only 0.3% Women of reproductive age have health insurance coverage.
- The most prevalent type of health insurance is private insurance and mutual or community organization
- Education, media exposure, good financial standing and financial inclusion increases the odds of having health insurance coverage.

## Introduction

Health insurance is a key mechanism for financing healthcare globally, aligning with the Sustainable Development Goals (SDGs) agenda for 2030.[1] Universal Health Coverage (UHC), central to SDG target 3.8, underscores equitable access to essential health services as a fundamental right, free from financial hardship. Despite these commitments, 2 billion people worldwide face significant financial challenges, with 1 billion incurring severe out-of-pocket expenditures, and 344 million pushed further into financial distress due to healthcare costs.[1] These persistent inequalities are exacerbated in low- and middle income countries (LMICs), where demographic shifts, including ageing populations and rising chronic disease burdens, strain the financial sustainability of health systems.[2,3]

Bangladesh exemplifies the financial barriers to healthcare in LMICs. Approximately 26% of households experience catastrophic health expenditures (CHE) during hospitalization annually, while out-of-pocket medical expenses accounted for 64.3% of the nation’s total health expenditures in 2015, amounting to USD 1.49 billion.[4] Alarmingly, less than 1% of the population benefits from social health protection, as per the government’s statistics. [5]

Women in reproductive age groups face unique barriers to affordable healthcare, particularly in LMICs. They experience higher health risks and often shoulder caregiving responsibilities, further exacerbating their vulnerability.[1] In Bangladesh, limited evidence exists on the determinants of health insurance coverage among women of reproductive age. This study seeks to address this gap by estimating the proportion of women of this age group with HIC and analysing factors influencing their access to it using data from the National Demographic and Health Survey.

## Methods

### Data overview

We used data from the latest round of the Bangladesh Demographic and Health Survey (BDHS) of 2022. The data can be found on request from the DHS website [https://dhsprogram.com/]. For the current analysis, we used data collected from women aged 15-49 years from selected households. In total, 19987 eligible women were considered for analysis. Details regarding the methodology of the BDHS 2022 can be found elsewhere.[6]

Operation definitions

### Outcome variables

*Health insurance coverage (HIC):* We considered HIC as the outcome variable in this study which had two categories. Women with any HIC at the time of survey were categorized as ‘yes’; otherwise ‘no’. The BDHS considered socialsecurity, employer-based insurance, mutual health organization orcommunity based insurance, privately purchased commercial insurance and other as HIC.[7]

### Independent variables

The independent variables considered for the study were age (in complete years), husband’s age (in complete years), residence (urban/rural), sex of the household head (male/female), bank or other institution account (no/yes), women’s level of education (no education/upto primary/upto secondary/post-secondary), husband’s level of education (upto primary/upto secondary/post secondary), wealth index (poorest/poor/middle/rich/richest), currently employed (no/yes), women empowerment (low/middle/high) and media exposure (poor/adequate).

After literature review, the independent variables were classified. The wealth index operates on the assumption that ownership of tangible assets, access to services, and the availability of amenities are indicative of a household’s economic status relative to others within the country.[8,9] This method offers an indirect yet effective means of estimating economic standing without the need for explicit income or expenditure data.[10]

The Survey-based Women’s Empowerment Index (SWPER) was introduced and validated using data from the Demographic and Health Surveys (DHS) across 34 African nations in 2017.[ll] It provides a standardized, individual-level tool for comparing women’s empowerment across countries and over time. The SWPER encompasses three dimensions of empowerment, capturing the resources and agency of partnered women, whether in marital or cohabiting unions.[11] Following the recommendations of Ewerling *et al*., participants were categorized into three categories of women empowerment: low, middle and high.

Mothers’ media exposure was assessed using three indicators: ‘reading newspapers,’ ‘watching television,’ and ‘listening to the radio.’ Engagement was scored as O for ‘not at all,’ 1 for ‘less than once a week,’ and 2 for ‘at least once a week.’[8] The total scores were categorized as 0-2 (‘poor’) and 3-6 (‘adequate’).[12]

### Data analysis

We analysed data using appropriate statistical methods. For continuous variables, we reported median (IQR); while categorical variables were reported using frequency and percentage. “Type of HIC” was presented via parliament chart. A Bivariate binomial logistic regression model was developed and unadjusted odds ratio (UOR) with their corresponding 95% confidence interval (Cl) were reported. Statistical analysis was conducted using jamovi (version 4.6.13). A p-value of <0.05 was considered statistically significant.

### Patient and public involvement

Patients and or publics were not directly involved in designing, conducting, interpreting or dissemination of the results of the study.

## Results

The total number of women aged 15-49 years who were included in this analysis was 19987. The median (IQR) age of the women was 32.0 (25.0-39.0) years and for their husband’s it was 39.0 (32.0-47.0) years. Most (64.9%) of them were rural residents, and only 14.5% were from families with female household heads. The highest proportion of women (45.7%) as well as their husbands (40.8%) reported upto secondary level of education. Only 0.3% women reported having HIC (Table 01).

**Table 1:**
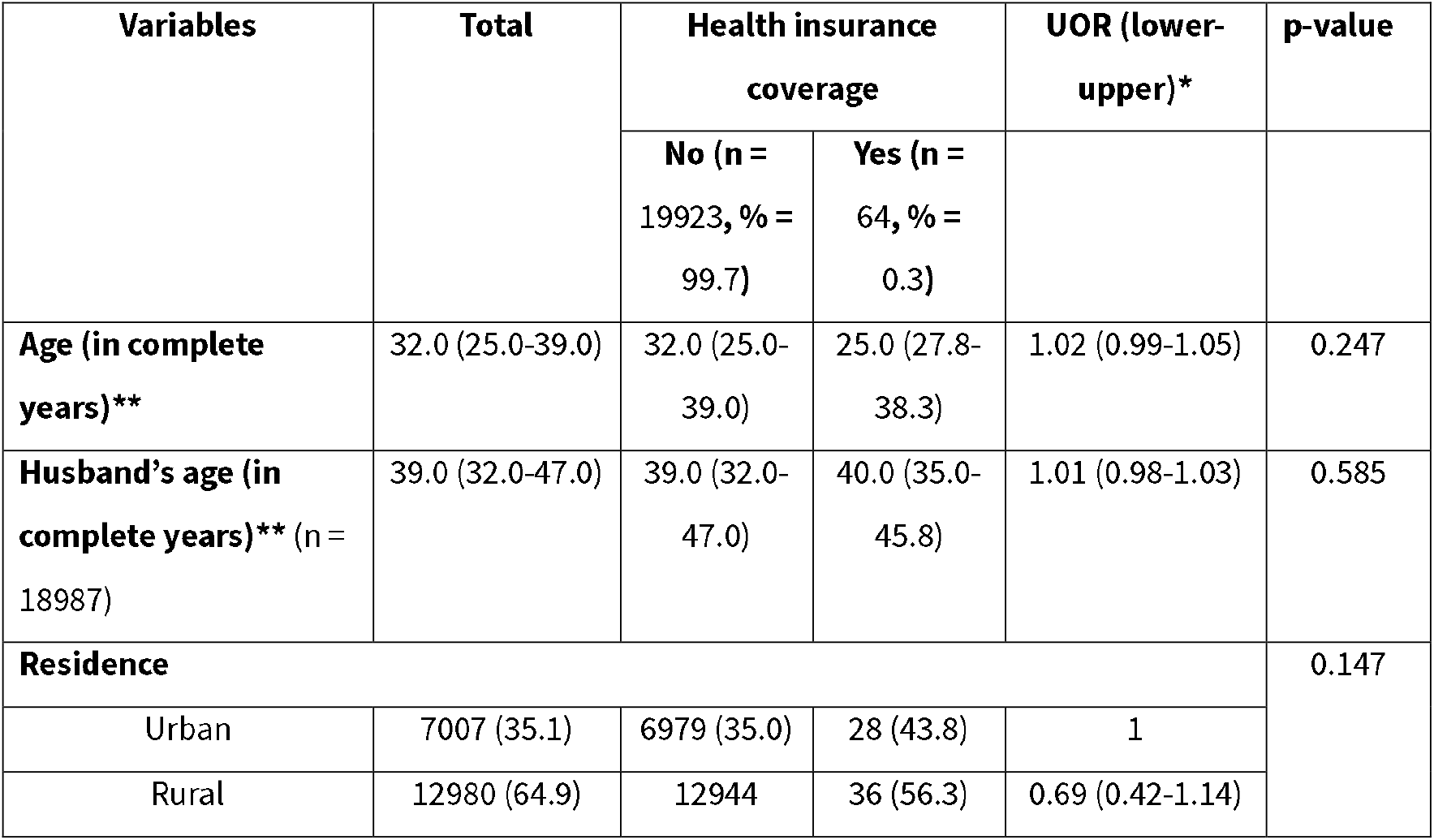
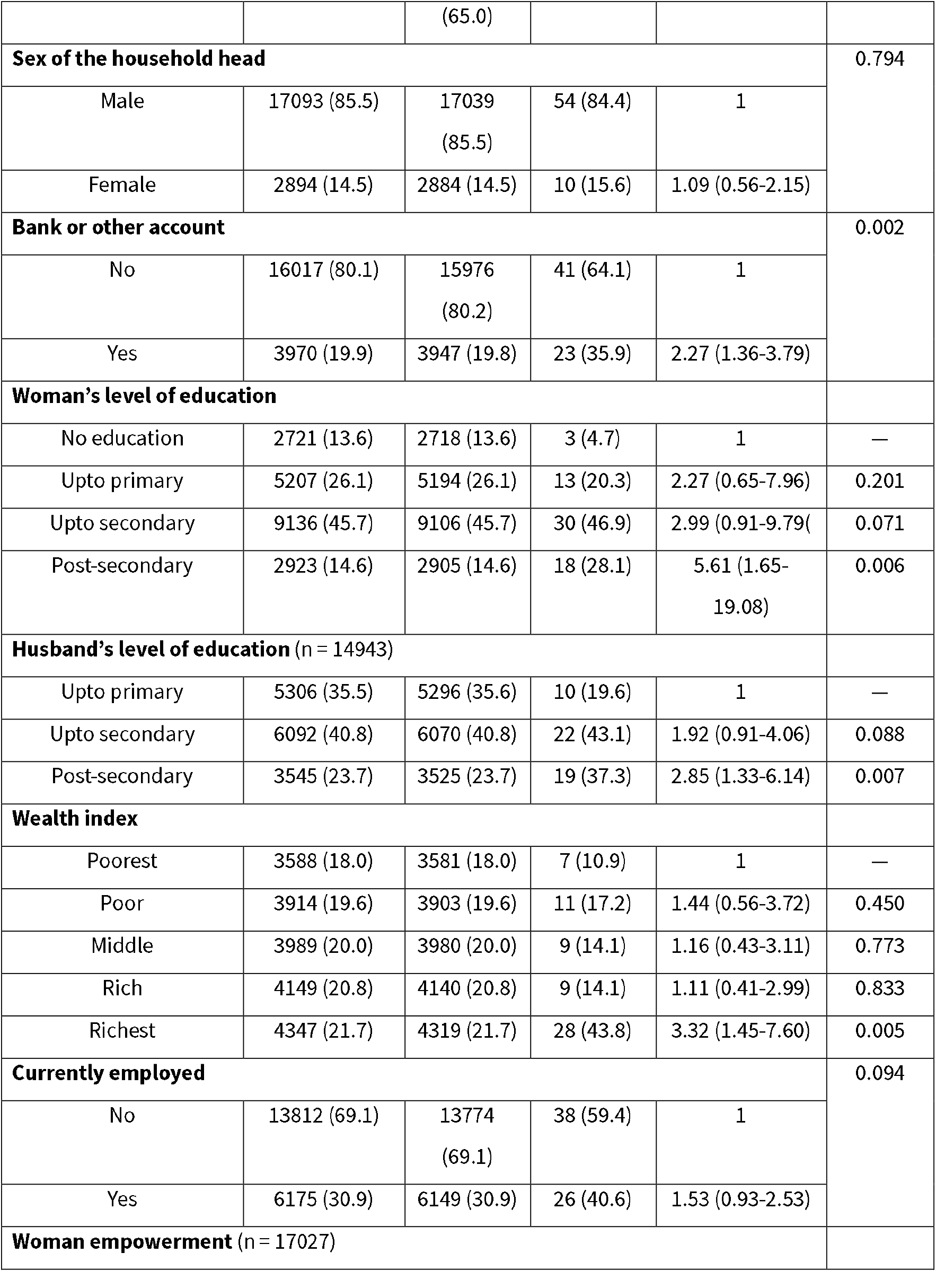
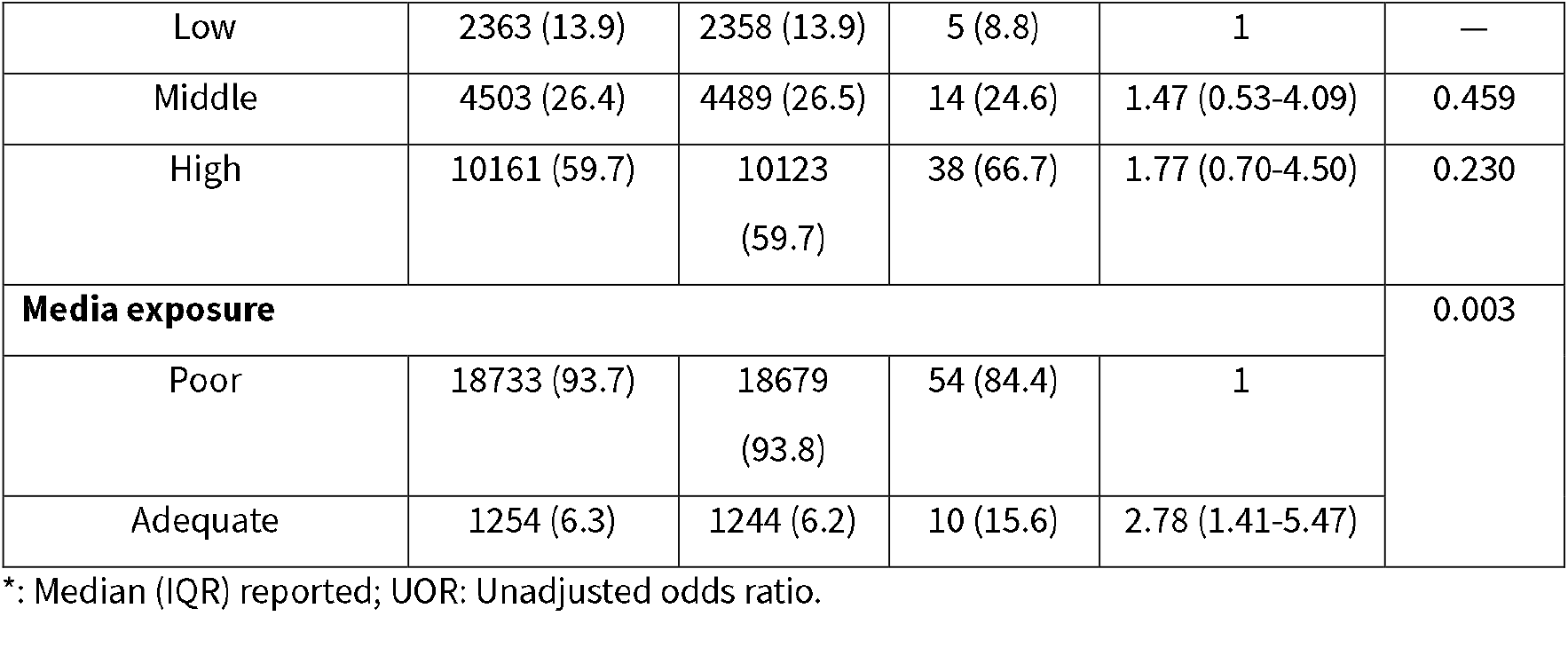
Socio-demographic characteristics of the Bangladeshi women.

Table 01 also reports that the odds of having HIC increased with having an account with a bank or other financial institution (UOR: 2.27; 95% Cl: 1.36-3.79; p-value=0.002). Similarly, increased odds of HIC were found among women with post-secondary education (UOR: 5.61; 95% Cl: 1.65-19.08; p-value=0.006), whose husbands had post-secondary education (UOR: 2.85; 95% Cl: 1.33-6.14; p-value=0.007), belonged to richest family (UOR: 3.32; 95% Cl: 1.45-7.60; p-value=0.005), and had adequate media exposure (UOR: 2.78; 95% Cl: 1.41-5.47; p-value=0.003).

Figure 01 illustrates the type of HI coverage found in the study. Private insurance coverage was reported by 35.4% participants, followed by coverage by mutual or community organization by 32.3% participants.

**Figure 1:**
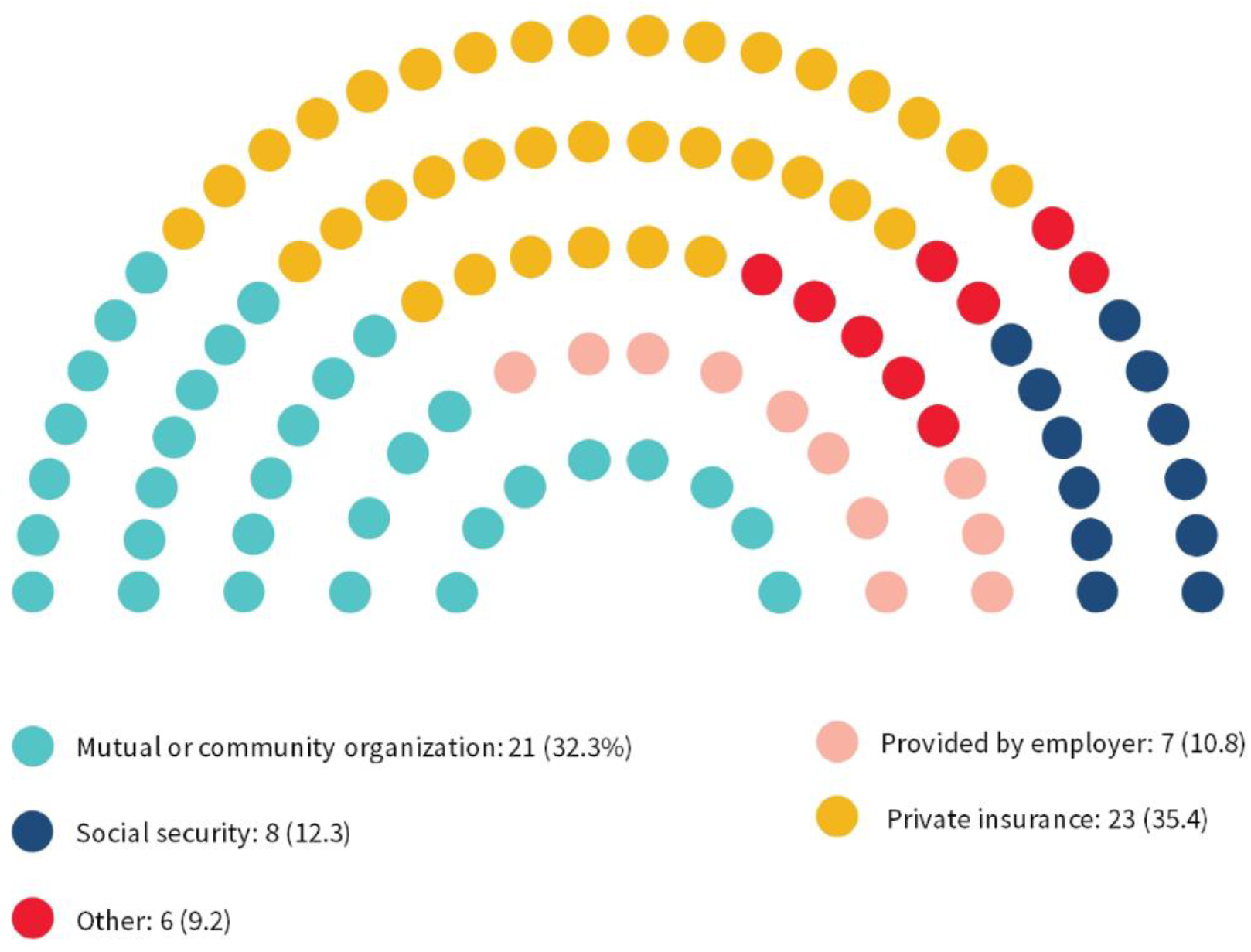
Type of health insurance (multiple response)

## Discussion

Health insurance is considered one of the key pathways which can help in achieving the SDG target 3.8 by promoting UHC.[l] Unfortunately, people from the LMICs, especially women in the reproductive age groups, are largely at a disadvantage as they have limited to no access to HIC.[1] In this study, we aimed to estimate the proportion of women of this age group with HIC and analyse factors influencing their access to it using data from the National Demographic and Health Survey. We found that only 0.3% of women have HIC, which corresponds to the national proportion of <l %.[5] The most common types of insurance were private insurance and mutual or community organizations. In Bangladesh, most insurance is provided by private companies, with only two state-owned insurers.[13] So, the preference for private insurance is understandable. Moreover, several community-based schemes operate at the local level, organized by NGOs and hospitals.[5] These schemes function as both insurers and service providers, offering integrated healthcare coverage, which explains the preference for coverage by mutual or community organizations. The low level of HIC prevalence can be attributed to the fact that the cost of premiums for private insurance is high and the virtual exclusion of the rural population and informal sector workers due to inaccessibility.[5]

Studies have suggested that individuals with higher levels of education are usually exposed to more health information, i.e., their health literacy increases.[14,15] As such, these individuals, especially women, tend to prefer HIC. Our study has also found a similar association with higher levels of education in the women or their husbands having higher odds of having HIC.

Socioeconomic status significantly influences health insurance ownership among women in developing countries. Studies indicate that women of reproductive age residing in wealthier households and advantaged neighbourhoods have higher chances of owning health insurance compared to their counterparts in poorer households and disadvantaged neighbourhoods.[14,16] We also found a similar association.

A study conducted in Ghana reports that women with greater exposure to media are more likely to uptake health insurance than their counterparts.[17] It has been suggested that radio and television are more accessible for women, and providing information via these media can promote HIC. Additionally, having an account with a bank or other financial organization is associated with a higher level of financial empowerment of women,[18] which tends to influence their decision towards HIC.

The strength of the study lies in the nationally representative data, which makes the findings generalizable. Additionally, this study explores the HIC among women of reproductive age, which provides much-needed insights into this much neglected area. However, there are also some limitations. The HI uptake is often influenced by a range of socio-economic factors, all of which are not included in the study. As it was a survey, no causal relationship could have been established.

## Conclusion

The current study highlights the extremely low prevalence of health insurance coverage among women of reproductive age in Bangladesh, with private and community-based insurance being the most common forms. The study also emphasises the need to promote both male and female education, at least up to a post-secondary level, to facilitate HIC. Additionally, targeted approaches should be made to bridge the inequality of coverage across variable wealth index quartiles and provide easy access to information related to HI via mass media.

## Data Availability

Data related to this manuscript are available upon request from the DHS website

https://dhsprogram.com/methodology/survey/survey-display-584.cfm

